# Trialling an Optimised social Groups intervention in services to Enhance social connecTedness and mental Health in vulnERable young people (TOGETHER): Study protocol for a feasibility randomised controlled trial

**DOI:** 10.1101/2023.07.04.23292217

**Authors:** Claire Vella, Clio Berry, Matthew J. Easterbrook, Anna-Marie Bibby-Jones, Daniel Michelson, Leanne Bogen-Johnston, David Fowler

**Affiliations:** University of Sussex, UK; Brighton and Sussex Medical School, UK; Sussex Partnership NHS Foundation Trust, UK; King’s College London, UK

**Author notes:** Corresponding author information: Claire Vella, School of Psychology, University of Sussex, Falmer, BN1 9QH, UK,. **Availability of data and materials:** No datasets were generated or analysed during the current study. All relevant data from this study will be made available upon study completion.

**Keywords:** Social connection, social groups intervention, mental health, young people, feasibility study

## Abstract

**Background:** Calls have been made to rethink the mental health support currently available for young people. This study aims to help re-focus and reduce the inaccessibility of mental health services by offering an adapted version of a theoretically-driven, evidence-based, guided psychosocial intervention known as ‘Groups 4 Health’ (G4H). To date, the G4H intervention has mainly been trialled in Australia, with promising positive effects on social connection, mental health and well-being. The present study examines the feasibility of running a randomised controlled trial when delivering the G4H intervention for young people in the UK.

**Methods:** The TOGETHER study is a feasibility randomised controlled trial of an adapted version of the G4H intervention. Participants are aged 16-25, currently experiencing mental health difficulties and recruited from mental health services. The target sample size is 30, with 15 in each trial arm. Participants are randomly allocated to either G4H plus treatment as usual, or treatment as usual alone. The primary outcomes of interest are the feasibility of recruitment, randomisation, data collection and retention to the study at 10 and 14 week follow up, as well as the acceptability, and accessibility of the study protocol and G4H intervention.

**Discussion:** The results of this study will indicate if further optimisation is required to improve the feasibility, acceptability and accessibility of the intervention and study protocol procedures as perceived by end users and practitioners. This offers a significant opportunity to support the local and national demand for accessible, innovative, and effective psychosocial youth mental health support.

**Trial registration:** ISRCTN registry (ISRCTN12505807). Registration date: 11/04/2022.

## Background

Severe and enduring mental health problems begin in adolescence, often emerging between the age of 15 and 25 years^1^. Despite a clear need to focus on young people’s mental health, concerns about the availability and adequacy of mental health support have risen considerably, with an increasing narrative from young people that their needs are not being met^2-4^. This is often particularly true for vulnerable young people, such as those with complex and severe mental health difficulties, unstable or inactive employment, and low socioeconomic status, who may disengage from, or may not meet the diagnostic entry thresholds for the standard NHS youth mental health services.

Calls have been made for the government to commit to a recovery plan for young people that rethinks the mental health support currently available^5^. Recommendations have been made to utilize the thousands of professionals working with young people who currently have limited or no training in psychological approaches^4^. Recommendations have also been made to implement practice innovations that directly address social isolation^6-7^. Research has consistently shown that the presence, severity and longevity of physical and mental health problems in young people is associated, and often preceded, by increased loneliness and diminished social interactions^4,8-9^. In contrast, social connection and multiple group memberships are associated with improvements in mental health symptoms^10^, well-being^11^, and resilience^12^.

Vulnerable young people have been associated with smaller social networks^13^ and the presence of social disability^14^. Assisting young people to engage in social relationships and activities, may help to prevent the onset and longevity of severe mental illness, as well as reduce the use of mental health services over time^14-16^. Addressing the underlying social causes of mental ill health is now also particularly pertinent following the prolonged social restrictions enforced to control the spread of the COVID-19 virus^17-18^. It is evident that young people with complex needs, i.e. co-present mental health and social difficulties, when they can access services, receive varying packages of support within a complex landscape of youth and mental health services^19^. The provision of interventions focused on social factors is variable and often poor. Services often aim to focus on diagnostic remission, symptom reduction, or a narrow promotion on paid employment. Such approaches do not always well match the typical heterogeneity of youth mental health presentations, nor necessarily what is most meaningful to young people themselves^20^. Services supporting youth mental health should aim to strengthen resilience in social and existential (i.e. life meaning) domains, and serve the higher-order processes of social participation and recovery^20^.

This study aims to address the urgent need to re-focus youth mental health support towards enhancing social connection. It also aims to help reduce the inaccessibility and burden on mental health services by offering an evidence-based psychosocial intervention, known as ‘Groups 4 Health’ (G4H)^21^, to clinical, health and community youth services. G4H is a modularised, guided psychosocial intervention that does not require extensive training to use. The intervention is built on the social identity approach to health, which proposes that positive social identities (identities based on beneficial group memberships) provide the psychological resources necessary to promote and maintain good physical and mental health^22^. G4H targets social connection and loneliness (through group-based belonging) with concomitant benefits for mental health and well-being^23-24^.

Trials so far have shown that the G4H intervention is associated with improvements in social connectedness, depression, anxiety, stress and life satisfaction for young people and adults from clinical, student and general population samples who are experiencing loneliness or low mood through an increase in positive social identities^21^. Trials have also indicated that loneliness is reduced considerably more when participants have received the G4H intervention, than compared to a matched no-treatment control group, a treatment as usual group, and group cognitive behavioural therapy^23-25^.

To date, the G4H intervention has mainly been trialled in Australia, using a group-based delivery format. The recruited samples also tend to be adults, who meet the criteria for clinical symptoms of depression. Only one registered study has focused on the G4H intervention in the UK, and this study similarly focuses on the adaptation of the intervention for adults accessing treatment for depression in secondary care services. Questions therefore remain about the transferability of the intervention to, not just young people in UK based services, but also vulnerable young people with complex mental health and support needs. Based on the research team’s extensive experience of working on trials with young people with complex social and mental health needs, as well as a recent consultation with a specialist children and young people’s mental health service, it is considered that the intervention may be more accessible to vulnerable young people when delivered individually. Further research is now needed to understand the feasibility, accessibility and acceptability of the intervention when delivered in this way. In the context of a complex youth mental health service landscape, there is a need to better understand what types of services could feasibly deliver a brief manualised social groups and connectedness intervention, and in what types of services this intervention may be effective. Moreover, the recommendations to utilise professionals with limited or no training in psychological approaches to augment youth mental health service provision^4^ does not necessarily mean that these professionals want to or will deliver the type of intervention being tested in this study. Thus, there is a need to explore the mechanisms and contextual factors that may affect the future uptake and implementation of the intervention, in both research and clinical contexts.

The present study will be the first known to examine the feasibility of delivering the G4H intervention with vulnerable young people in UK based community, health and youth mental health services. This study will also examine the feasibility parameters related to the design of a randomised controlled trial, in such UK based services. The results will indicate if further optimisation or adaptations are required to improve the feasibility, acceptability and accessibility of the intervention and study protocol procedures, as perceived by end users and (actual and potential) intervention providers. Findings will also provide valuable information from actual and potential intervention providers regarding their perspectives around intervention implementation. This will inform the overarching intervention implementation strategy, ensuring too that future research studies accurately reflect this intended end deployment. This study offers a significant opportunity to better meet the local and national demand for accessible, innovative, and effective youth mental health support.

### Aims

The project aims to evaluate the feasibility, acceptability and accessibility of a randomised controlled trial delivering an adapted social connection intervention (Groups 4 Health; G4H) to young adult service-users who are currently experiencing mental health difficulties.

#### Primary research questions

1) Is it feasible to conduct a randomised controlled trial for the G4H intervention when delivered to young service-users accessing community, health and/or youth mental health services?
2) Is the G4H intervention feasible to deliver to young service-users involved in community, health and/or youth mental health services?
3) Is the G4H intervention safe and acceptable according to both the young service-users receiving the intervention, and the practitioners delivering the intervention?
4) What changes are indicated to improve the safety, acceptability, accessibility and feasibility of the intervention?

#### Secondary research questions

5) What is the most suitable primary outcome for a future larger trial of the G4H intervention?
6) What are the attitudes, experiences and contextual factors relevant to the implementation of an intervention for social connectedness by practitioners’ working with young people with mental health problems?
7) What are young people’s experiences and perceptions of the benefits and harm of identification with online-based social groups?

## Methods

### Trial design

TOGETHER is a single-blind, parallel-group feasibility randomised controlled trial, and a multi-site single time-point online practitioner implementation survey. Participants aged 16-25 who are currently experiencing mental health difficulties will be independently randomised to receive either an adapted version of the G4H intervention alongside treatment as usual, or treatment as usual only. The trial will run in the context of community, health and/or youth mental health service settings in the South East of England. The practitioner implementation survey will run in service settings nationally. The CONSORT (Consolidated Standards of Reporting Trials) Statement and CONSORT Checklists will be used to report the trial. For this protocol, the SPIRIT (Standard Protocol Items: Recommendations for Interventional Trials) Figure and Checklist are provided (See Supporting Information file S1).

### Participants

The study will recruit trial young adult participants, intervention providers and practitioner survey respondents. See Supporting Information file 2 for the eligibility criteria to take part as an intervention provider and practitioner survey respondent. The trial aims to recruit 30 young adult participants, providing 15 participants in each trial arm.

#### Inclusion criteria

1. Aged 16 – 25 years old
2. Accessing a community, health and/or youth mental health service involved in the study
3. Be experiencing current mental health difficulties (operationalised by a rating of 60 or below on the Global Assessment Scale [GAS])
4. Able to read, write and speak in English/ OR are non-English speaking but have access to an interpreter; to the degree they can give informed consent, are able to fully understand and participate in both the assessment questions and intervention content

#### Exclusion criteria

1. Be at immediate and serious risk to self or others (assessed at the point of referral/eligibility review)
2. Be currently participating, or be confirmed to participate in another interventional research study in which they are receiving an intervention that targets social isolation or utilises psychological therapy
3. Be expected to be discharged, or be known to be unable to seek support from the referring service, in the 16 weeks following a referral to the trial being made (14-week research involvement + 2 weeks allowing for any missed/rearranged meetings)

### Intervention

#### G4H

The intervention is G4H plus TAU. G4H is a theoretically driven, evidence-based, manualised intervention designed to improve social connection and well-being through facilitating integration to social groups^26^. The intervention is delivered via five sessions, each taking approximately 60 to 90 minutes. The first four sessions are delivered weekly, and the final session is delivered one month later. The G4H sessions are supplemented with a workbook, offering a summary of the main learning points and space to complete activities. The five sessions aim to: raise awareness of the value and benefits of groups for health; develop a social map to identify existing connections and possible areas for social growth; train skills to maintain and use existing networks, and/or reconnect with valued groups; explore if existing connections reflect the important aspects of the self, and create an action plan; reinforce key messages and troubleshoot any problems experienced with implementing the developed social action plan.

The G4H intervention is delivered by trained intervention providers who are either working in the services who have agreed to be involved in the trial or are associated members of the research team with relevant experience. Following consultation with practitioners and young people (see PPI section), the trial offers an adapted version of G4H (see Patient and Public Involvement section). For this study, the sessions are typically delivered in a 1:1 format, with the option to take part in sessions as a group. The intervention sessions are held either online or in-person; allowing services to follow their usual policies and/or procedures for contact with service-users.

#### TAU

The control comparator is treatment as usual. This consists of any support, medication or interventions that participants may be receiving from any statutory or non-statutory public services and/or private services.

#### Intervention training & adherence

Those who have consented to be intervention providers in the study are invited to a group training session delivered by a member of the Groups 4 Health team. The training session may be held in-person or online using videoconferencing facilities. Intervention providers are provided with a copy of the intervention manual and workbook and asked to follow the detailed intervention manual where possible. For each delivered session of the intervention, intervention providers are asked to complete an intervention adherence form. Questions relating to perceived fidelity to the intervention, and perceived barriers to maintain fidelity to the intervention are asked in a semi-structured interview with intervention providers.

### Outcomes

#### Primary outcomes

The primary outcomes of interest are:

1. Number of potential service-user participants referred
2. Number of intervention providers who consent to take part
3. Number and proportion of referred potential service-user participants who consent to take part in the study
4. Number and proportion of referred potential service-user participants found to be eligible
5. Number and proportion of consenting eligible participants who are retained in the study at post-intervention (10 week follow up) and until the last time point (14-week follow up)
6. Number and proportion of survey measures completed by each participant for each time-point
7. Number and proportion of consenting eligible service-user participants who take part in all five G4H sessions
8. Number and proportion of intervention adherence components completed for each session and across the whole G4H intervention
9. Number and nature of adverse events experienced by study participants
10. Number and proportion of breaks in blinding

The key feasibility parameters for the study include:

- ≥ 50% of referred potential participants will be found to be eligible
- ≥ 80% of consenting eligible participants will be retained in the study at each follow up time point
- ≥ 80% data completeness
- ≥ 80% of participants allocated to receive G4H will complete at least four sessions
- ≥ 80% of the core G4H intervention items will be delivered

#### Secondary outcome

The secondary outcome is defining the primary outcome for a future larger trial of the G4H intervention.

#### Participant perspectives of feasibility, acceptability, accessibility, and safety

To assess the feasibility, acceptability, accessibility and safety of the G4H intervention and study protocol from a participant perspective, qualitative data are collected from the young people and intervention providers involved in this study. To further understand the attitudes, experiences and contextual factors related to the implementation of an intervention for social connectedness, both quantitative and qualitative data drawing on Normalisation Process Theory^27^ is also collected via the study practitioner national survey.

### Recruitment and procedure

The trial will run in the context of community, health and/or youth mental health service settings in the South East of England. The study aims to include: the Sussex Partnership NHS Foundation Trust Children and Adolescent’s Assertive Outreach Team; a Sussex, Kent or Surrey based Primary Care service; a Sussex-based YMCA youth advice centre or residential service. The practitioner implementation survey will be open to NHS and non-NHS services across the UK.

To recruit young adult participants, all organisations who have agreed to be involved in the trial are provided with the young adult-focused participant information sheet, study information video (with video transcript), study poster, referral information sheet and referral form. The involved organisations are asked to discuss the study opportunity and/or share the study documents with potential eligible young people. Potential participants who are interested and provide verbal permission to be contacted by the researcher, are referred by the involved organisation. Researchers then invite potential participants to provide informed consent and to be screened for study eligibility. Eligible participants are then invited to complete the baseline assessment and, upon doing so, are randomised. All randomised participants are invited to complete two post-allocation follow up assessments at 10 and 14 weeks. Participants receive £20 in vouchers (£60 in total) for each assessment time point. Please refer to Fig. 1 (SPIRIT figure) for the schedule of enrolment, allocation, intervention and assessments. For information about the recruitment and consent process for intervention providers and practitioner survey respondents, see Supporting Information file 2.

**Fig. 1.**
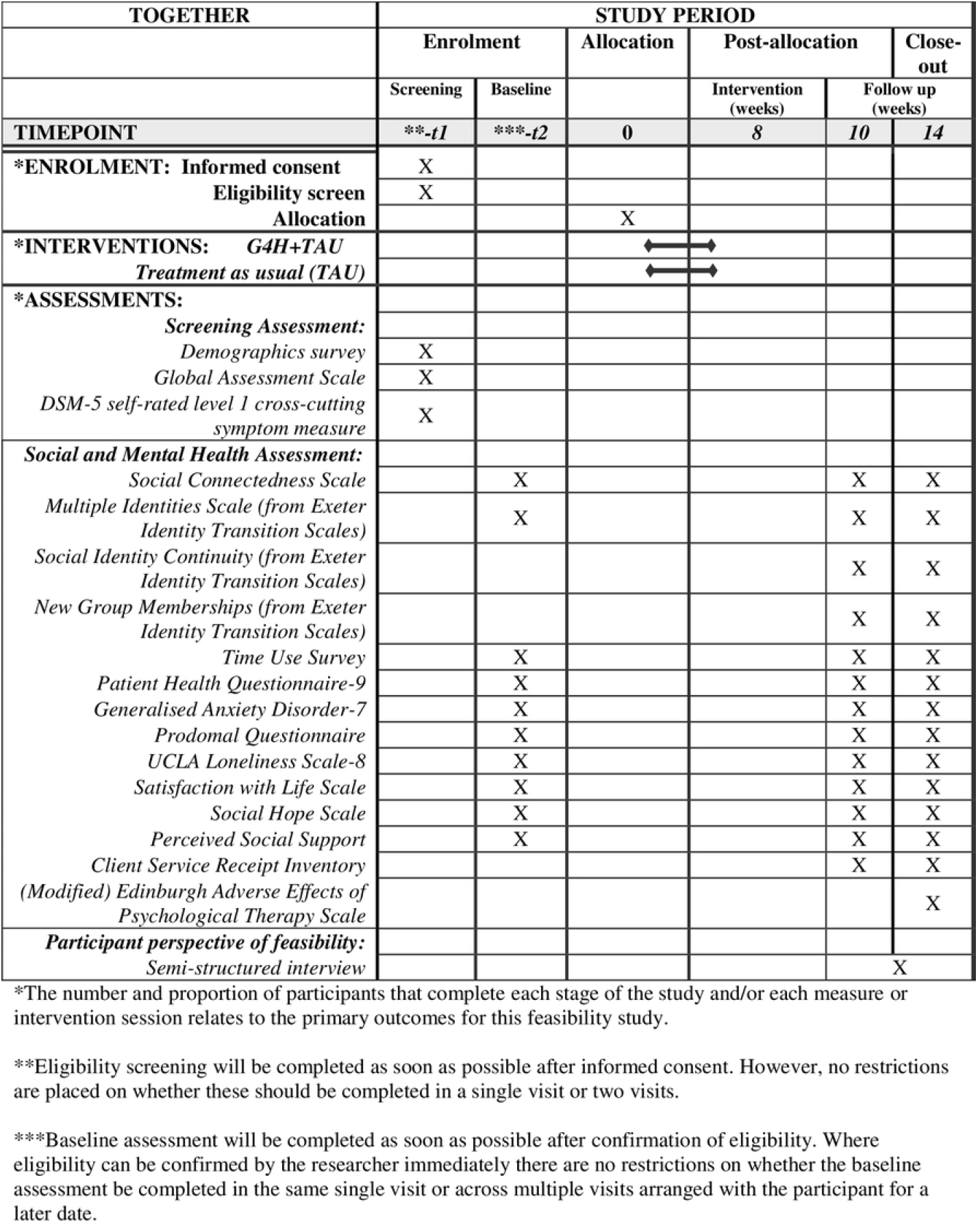
Standard Protocol Items: Recommendations for Interventional Trials (SPIRIT); Schedule of enrolment, interventions and assessments.

All trial participants are invited to complete a semi-structured interview (either post 10 week follow up assessment or post intervention delivery, as relevant to the type of participant). Verbal consent to be audio-recorded is obtained. Across the trial, all personal information and research data is collected online, on a day and a time suitable for the participant; either independently or with a researcher in-person or via telephone or email. If required, a member of the research team may read study documents to the individual, and/or record participant responses on their behalf (with participant permission).

### Ethical approval and consent to participate

Ethical approval for the study was provided by the Health Research Authority, Berkshire Research Ethics Committee (Reference: 22/SC/0040). Participants are required to provide written informed consent to take part and are provided with the relevant participant information sheet at least 24 hours prior.

### Adverse events

All adverse events will be categorised and recorded from the point of randomisation until completion of the final follow up assessment for young adult participants, and from the point of informed consent until post-intervention interview for intervention providers. All serious adverse events, and suspected, unexpected, serious adverse reactions (SUSARs) will be reported to study sponsor and NHS research ethics committee as appropriate.

### Randomisation & Blinding

The randomisation procedure and allocation sequence is set up by an independent member of the research team and will be masked from all other study team members. Randomisation, stratified by trial site, is completed using the Sealed Envelope online service^28^. Treatment allocation is emailed to an unblind trial coordinator who contacts each trial participant (via phone, email or preferred contact method) to inform them of the group they have been allocated to. The referring service, participant’s GP, and secondary care mental health service (where relevant) are informed of the participant’s informed consent to the study and allocated treatment group. For participants allocated to receive G4H alongside treatment as usual, an assigned intervention provider is advised to set-up arrangements to start delivering the intervention as soon as possible. Research workers responsible for data collection from each follow up assessment battery are kept blind (where possible) to treatment allocation to facilitate unbiased, and objective assessments. All trial participants, intervention providers and referring services are asked not to reveal the allocation group to the assessor. Trial allocation is stored in a file not accessible to blinded members of the team. Breaks in blinding are monitored and recorded. Qualitative interviews are completed by an unblind member of the team.

### Patient and Public Involvement

This protocol is informed by the feedback received from stakeholders and young people aged 18-25 with lived experience of mental health problems. The concept of the study, involving an intervention that targets social connectedness for young people’s mental health, was presented and supported as an interesting and useful opportunity by experts by experience at a Young People’s involvement in Digital Mental Health research group event organised by the University of Nottingham and Emerging Minds research network, as well as by stakeholders from Sussex Partnership NHS Foundation Trust (notably the chief medical officer, and practitioners from a specialist youth mental service). The research team engaged in a process of co-production to optimise the chosen intervention to overcome any perceived immediate accessibility issues for our target population. Key adaptations were made following informal interviews with four young adults with lived experience (paid for their time) and a focus group with five practitioners from a specialist youth mental health service. These include: offering the delivery of the G4H intervention in a 1:1 format; flexibility to deliver online or in-person; adapting content language; and option to use modified scales for session activities.

### Sample Size

The trial aims to recruit 30 young adult participants, providing 15 participants in each trial arm. The target sample size exceeds the recommended minimum sample size for feasibility studies^29^, and a power calculation was not undertaken as the study focus is on feasibility goals and not the statistical significance of treatment effects^30^. Similarly, the sample size of intervention providers is not pre-determined and is dependent on the number of eligible practitioners in each of the services involved in this study. This feasibility study aims to monitor the number of intervention providers who consent to take part to inform the design of a larger RCT study. The study will also recruit a minimum of 100 survey respondents from UK services involved in supporting young people aged 16-25 to complete the practitioner implementation survey.

### Data Management

All personal and research data (online, electronic or paper-based) is processed in accordance with the General Data Protection Regulation and Data Protection Act (2018). Data is stored confidentially and securely with log in/password protection and/or in locked filing cabinets on NHS premises. All research data is pseudo-anonymised with a unique participant identification number. Participant personal information is stored separately in a password protected file. Only fully anonymised data is available for the use of other genuine researchers or as a part of publication transparency. Participant consent for anonymised data to be shared outside of the research team is provided during the informed consent process.

### Analysis

This study collects both quantitative and qualitative research data. Quantitative data will be evaluated using descriptive statistics (count, proportions, mean, median, standard deviation, IQR, range & reliable change analysis) as appropriate. Participant flow through the study will be reported in line with the CONSORT 2010 statement - extension to randomised pilot and feasibility trials^31^. Qualitative data transcribed verbatim into text files will be analysed using framework analysis. Missing data will be handled according to best practice guidelines^32^ and will be evaluated with respect to the amount and nature of the missing data, and any substantive patterns related to the missing data. The level of missing data will be reported.

## Discussion

There is an urgent need to reduce the inaccessibility and burden on mental health services. Services often aim to focus on diagnostic remission, symptom reduction, or a narrow promotion on paid employment, and the provision of interventions focused on social factors is variable and often poor. Evidence suggests that assisting young people to enhance their sense of social connectedness and to engage in social relationships and activities may help to prevent the onset and longevity of severe mental illness, as well as reduce the use of mental health services over time. This study will be the first known to examine the feasibility, accessibility and acceptability of delivering a short, evidence-based psychosocial intervention, known as ‘Groups 4 Health’ (G4H), for vulnerable young people in UK based community, health and youth mental health services. The study seeks to uniquely investigate the transferability of the intervention to young people in UK-based services, and in particular the accessibility of the intervention for vulnerable young people when delivered individually. The mechanisms and contextual factors that may affect the future uptake and implementation of the intervention in research and clinical contexts will also be explored. Such research offers a significant opportunity to better understand and meet the local and national demand for accessible, innovative, and effective social-based youth mental health support. The study team plan to work with key stakeholders to co-produce videos and a lay summary of the study results to disseminate broadly through clinical, research, academic and public engagement activities. The findings will also be published in peer-reviewed journals and presented at local and international conferences.

### Protocol changes

The study sponsor and NHS Research Ethics Committee are notified of amendments to the study protocol. Amendments are not enforced until the relevant approvals have been received. All amendments made to the study protocol since commencement of the trial are shown in Table 1.

**Table 1.** Amendments to protocol.

| Area | Change |
| --- | --- |
| Design | Inclusion of a multi-site practitioner implementation survey; reduced service-user participant recruitment to a total of 30 participants; inclusion of additional safeguarding criteria and procedures |
| Data Collection | Changes to assessment battery: Addition of modified Client Service Receipt Inventory to 10 week and 14 week follow up |
| Recruitment Procedure | Use of a study information video |
| Trial Timeline | 3-month study extension; 2-month study extension |

### Trial status

Recruitment of participants commenced in May 2022 and will be open until the end of June 2023. Delivery of the intervention will continue until the end of August 2023, and the collection of research data will continue until early October 2023.

## Data Availability

No datasets were generated or analysed during the current study. All relevant data from this study will be made available upon study completion.

## Authors’ contributions

CV took responsibility for the main drafting of the protocol and manuscript. All authors made substantial contributions to the conception and design of the study and the development of the protocol. All authors read and approved the final manuscript.

## Supporting Information

Supporting Information file S1 – SPIRIT Checklist

Supporting Information file S2 – Eligibility criteria and recruitment processes for intervention providers and practitioner survey respondents.

